# A Logistic Curve in the SIR Model and Its Application to Deaths by COVID-19 in Japan

**DOI:** 10.1101/2020.06.25.20139865

**Authors:** Takesi Saito, Kazuyasu Shigemoto

**Affiliations:** Department of Physics, Kwansei Gakuin University, Sanda 669-1337, Japan; Tezukayama University, Nara 631-8501, Japan

## Abstract

Approximate solutions of SIR equations are given, based on a logistic growth curve in the Biology. These solutions are applied to fix the basic reproduction number *α* and the removed ratio *c*, especially from data of accumulated number of deaths in Japan COVID-19. We then discuss the end of the epidemic. These logistic curve results are compared with the exact results of the SIR model.

## 1 Introduction

The SIR model [1] in the theory of infection is powerful to analyze an epidemic about how it spreads and how it ends [2–8]. The SIR model is composed of three equations for *S, I* and *R*, where they are numbers for susceptibles, infectives and removed, respectively. Three equations can be solved completely by means of MATHEMATICA, if two parameters *α* and *c* are given, where *α* is the basic reproduction number and *c* the removed ratio.

In Sec. 2 we would like to summarize some exact solutions of the SIR equations. These exact solutions are applied to COVID-19 in Japan. Here, our policy is a little use of data of cases. In Sec. 3 we propose approximate solutions of SIR equations, based on the logistic growth curve in the Biology [9]. These approximate solutions have simple forms, so that they are very useful to discuss an epidemic. The final section is devoted to concluding remarks. The logistic approach is compared with exact solutions of the SIR model to our epidemic.

## 2 The SIR model in the theory of infection

Equations of the SIR model are given by

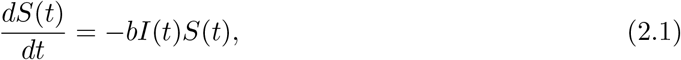

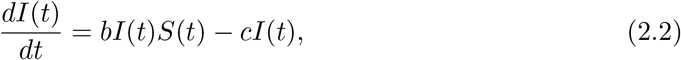

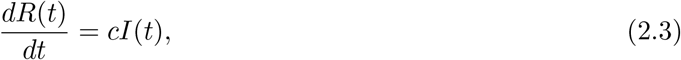

where *S, I* and *R* are numbers for susceptibles, infectives and removed, respectively, *b* the infection ratio and *c* the removed ratio. Our aim is to propose an approximate solutions of these equations, based on a logistic growth curve in the Biology.

We first summarize some of exact solutions of the SIR model. From Eq(2.1) and Eq.(2.3) we get *dS*/*dR* = −*αS*, (*α* = *b*/*c*), which is integrated to be

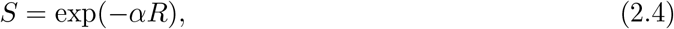

where *α* stands for the basic reproduction number. In the same way, from Eq.(2.2) and Eq.(2.4), we have *dI*/*dR* = *αS* − 1 = *α* exp(−*αR*) − 1, which is integrated to be

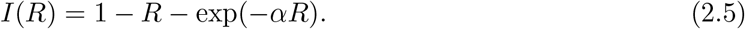

The solutions Eq.(2.4) and Eq.(2.5) satisfy boundary conditions *S* = 1 and *I* = 0 at *R* = 0. We normalize the total number to be unity, i.e., *S* + *I* + *R* = 1. Since *I*(*t*) is 0 when *t ⟶* ∞, we have 1 − *R*(∞) − exp [−*αR*(∞)] = 0 from Eq.(2.5). Hence, it follows a useful formula

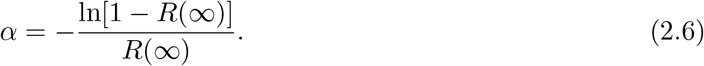

Some exact formulas at the peak *t* = *T* are summarized as follows:

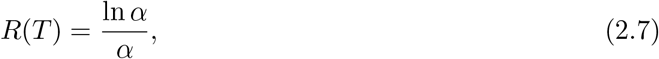

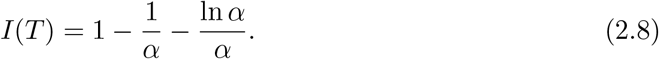

The first one is derived as follows: Since the peak point is given by *dI*/*dt* = *c*(*αS* − 1)*I* = 0 at *t* = *T*, we get *S*(*T*) = exp[−*αR*(*T*)] = 1/*α*. This proves Eq.(2.7). The second equation (2.8) directly follows by *I*(*T*) = 1 − *R*(*T*) − *S*(*T*) = 1 − ln *α*/*α* − 1/*α*.

Now let us consider an application of the above exact results in the SIR model to COVID-19 in Japan. In order to fix *T, α* and *c*, we use the data of deaths [10]. At May 2nd, the accumulated number of deaths *D* takes 492 and the new increased number of deaths Δ*.D*(*t*)/Δ*.t* takes the maximum value 34. Then we find *T* to be May 2nd. Here, our policy is a little use of data of cases. Then we connect *D*(*t*) with *R*(*t*) at *t* = *T* by the formula *D*(*t*) = *rR*(*t*), where *r* is the death rate, so that

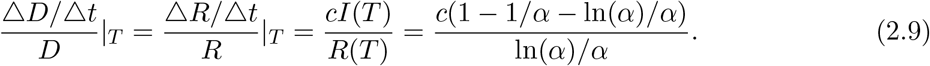

Since Δ*.D*(*t*)/Δ*.t* is fluctuating, we take 5-day average from April 30th to May 4th. The 5-day values for Δ*.D*(*t*)/Δ*.t* are (April 30th: 17), (May 1st: 26), (May 2nd: 34), (May 3rd :18), (May 4th: 11), and the 5-day average around May 2nd gives Δ*.D*/Δ*.t*|_5-day average_ = 21.2. By using *D*(*T*) = 492, we have (Δ*.D*/Δ*.t*)/*D*|_*T*_ = 21.2/492 = 0.043.

Next, let us fix *c* from the formula *c* = (Δ*.R*(*t*)/Δ*.t*)/*I*(*t*). We take the total average from Feb. 16 to June 9, in order to compensate the fluctuation. Thus we get *c* = (Δ*.R*(*t*)/Δ*.t*)/*I*(*t*)|_total average_ = 0.041, which means that almost all cases are rejected approximately after 25 days. Substituting *c* = 0.041 into Eq.(2.9), we have an equation for *α*

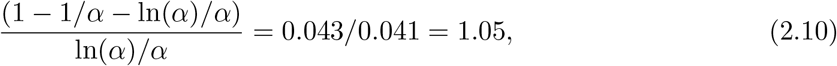

which gives a solution *α*_exact_ = 3.66.

To sum up, we have exact values of parameters: *T* =May 2nd, *α*_exact_ = 3.66 and *c* = 0.041. We then draw curves of *S, I* and *R* by means of MATHEMATICA in Fig. 1.

**Figure 1:**
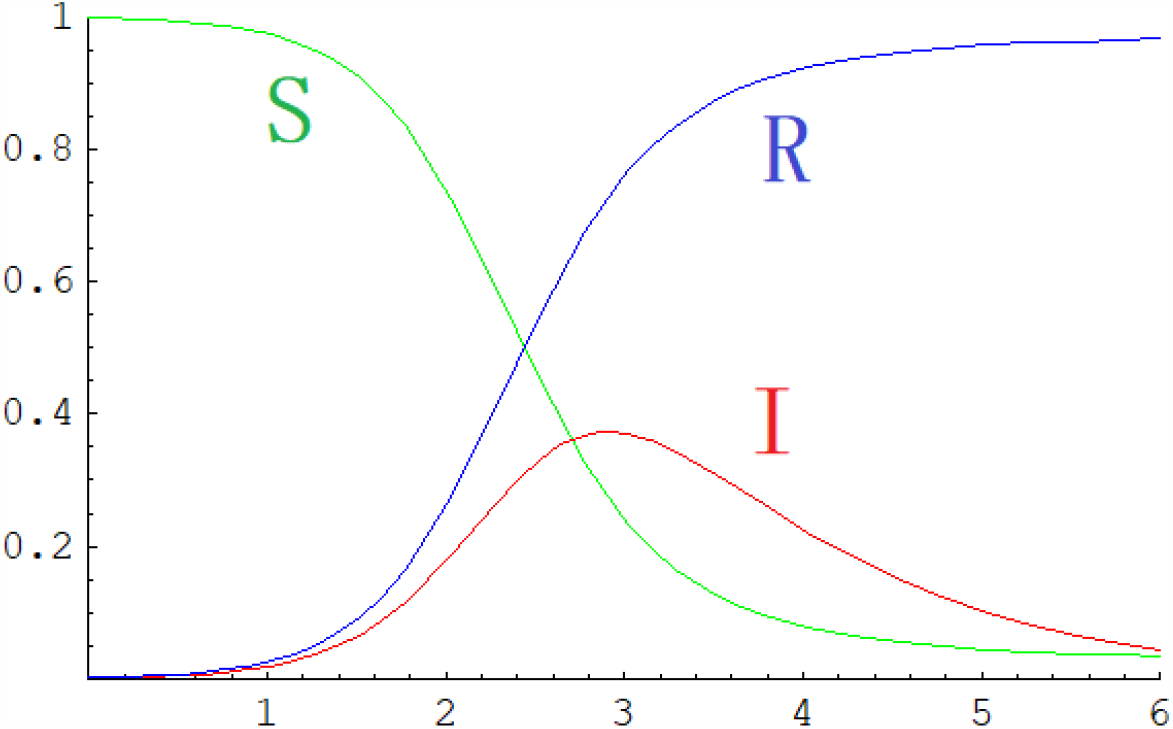
Graph of *S, I* and *R* for α = 3.66 and *c* = 0.041.

## 3 A logistic curve from the SIR model

Let us consider approximate formulas for SIR’s functions. The third equation Eq.(2.3) can be written as

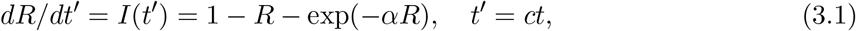

with *t* the true time. Let us expand the exponential factor in the second order of *x* = *αR*,

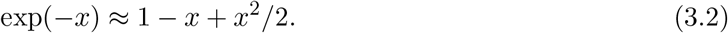

Then we have

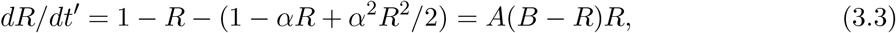

with *A* = *α*^2^/2, *B* = 2(*α* − 1)/*α*^2^. This equation is a type of the logistic growth curve in the Biology [9], easily solved as

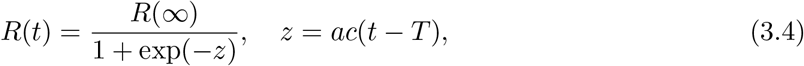

where *AB* = *α* − 1 = *a* and *B* = *R*(∞) = 2(*α* − 1)/*α*^2^ = 2*R*(*T*). Inserting *R*(*t*) into

*I*(*t*) = (1/*c*)(*dR*(*t*)/*dt*), we have

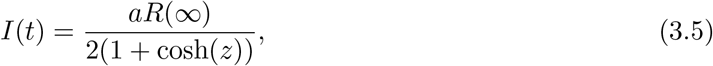

where the peak value of *I*(*t*) is given by *I*(*T*) = *aR*(∞)/4 at *t* = *T* .

Here we have useful formulas

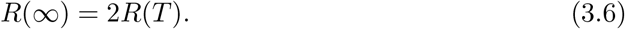

In the following, we consider an application of the logistic curve to COVID-19 in Japan.

By using Eq.(2.9), Eq.(3.4) and Eq.(3.5), we have

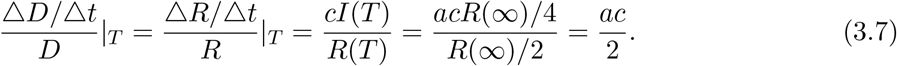

According to the data of accumulated number of deaths, we have (Δ*.D*/Δ*.t*)/*D*|_*T*_ = 21.2/492 = 0.043, which gives *α*_appr_ = *a* + 1 = 3.10. In this way, we have fixed parameters in the logistic curve: *T* =May 2nd, *α*_appr_ = 3.10 and *c* = 0.041. The value *α*_appr_ = 3.10 should be compared with *α*_exact_ = 3.66. The error of our approximation for *α* is about 15%.

## 4 Concluding remarks

We have proposed logistic formulas in the theory of infection, which are approximate formulas driven from the SIR model. These formulas appear to be simple forms, therefore, very useful to analyze an epidemic. In logistic formulas we have determined the basic reproduction number *α*_appr_ = 3.10 with the removed ratio *c* = 0.041, from data of the accumulated number of deaths in Japan COVID-19. One can see *D*(∞) = 984 from the approximate formula *D*(∞) = 2*D*(*T*) = 2 × 492, that is, the final accumulated number of deaths is 984.

We have also fixed exactly *α* and *c*, in the SIR model for Japan COVID-19. Once having the basic reproduction number *α*_exact_ = 3.66 and the removed ratio *c* = 0.041, we can draw curves of *S, I* and *R* by means of MATHEMATICA in Fig. 1.

The peak day of Δ*.D*(*t*)/Δ*.t* is *T* =May 2nd from the data, which is the 77th day from Feb.15. Since Δ*.D*(*t*)/Δ*.t* is almost left-right symmetric with respect to the peak day, the epidemic ends on 77 days after May 2nd, a middle of July.

Finally if we compare *α*_appr_ = 3.10 with *α*_exact_ = 3.66, the error of our approximation for *α* is about 15%.

## Data Availability

“Summary of the New Coronavirus Infection ”(in Japanese), https://hazard.yahoo.co.jp/article/20200207; “Status of the Domestic New Coronavirus Infection”(in Japanese), https://toyokeizai.net/sp/visual/tko/covid19/

